# Assessing the role of vascular risk factors in dementia: Mendelian randomization meta-analysis and comparison with observational estimates

**DOI:** 10.1101/2022.02.23.22271334

**Authors:** L Lee, R. M. Walker, W. N. Whiteley

**Affiliations:** Medical Student, University of Edinburgh, UK; Statistical Epidemiologist, University of Edinburgh, UK; Reader in Neurology and Consultant Neurologist, University of Edinburgh, UK

## Abstract

**Importance:** Although observational studies demonstrate that higher levels of vascular risk factors are associated with an increased risk of dementia, these associations might be explained by confounding or other biases. Mendelian randomization (MR) uses genetic instruments to test causal relationships in observational data.

**Objective:** To determine if genetically predicted modifiable risk factors (type 2 diabetes mellitus, low density lipoprotein cholesterol, high density lipoprotein cholesterol, total cholesterol, triglycerides, systolic blood pressure, diastolic blood pressure, body mass index, and circulating glucose) are associated with dementia by meta-analysing published MR studies. Secondary objectives were to identify heterogeneity in effect estimates across primary MR studies and to compare meta-analysis results with observational studies.

**Data sources:** MR studies identified by systematic search of Web of Science, OVID and Scopus.

**Study selection:** Primary MR studies investigating the modifiable risk factors of interest. Only one study from each cohort per risk factor was included. A quality assessment tool was developed to primarily assess the three assumptions of MR for each MR study.

**Data extraction and synthesis:** Data were extracted on study characteristics, exposure and outcome, effect estimates per unit increase, and measures of variation. Effect estimates were pooled to generate an overall estimate, I^2^ and Cochrane Q values using fixed-effect model.

**Main outcomes and measures:** Odds ratio (OR) of developing dementia per standardized unit increase in the risk factor of interest.

**Results:** We screened 5211 studies and included 12 primary MR studies after applying inclusion and exclusion criteria. Higher genetically predicted body mass index was associated with a higher odds of dementia (OR 1.03 [1.01, 1.05] per 5 kg/m^2^ increase, one study, p = 0.00285). Overall estimates from MR studies showed a smaller number of associations than those from meta-analyses of observational studies.

**Conclusion and relevance:** Genetically predicted body mass index was associated with an increase in risk of dementia.

**Key points:** *Question:* Are genetically predicted modifiable risk factors associated with dementia?

*Findings:* Genetically predicted higher body mass index was associated with a higher odds of dementia. No evidence was found to support an association between genetically predicted type 2 diabetes mellitus, low density lipoprotein (LDL) cholesterol, high density lipoprotein (HDL) cholesterol, total cholesterol, triglycerides, systolic blood pressure, diastolic blood pressure, plasma glucose and dementia risk.

*Meaning:* Many modifiable risk factors associated with dementia in observational studies may not play a causative role.

## Introduction

Higher measured mid-life blood pressure, mid-life and late-life hyperlipidaemia, mid-life obesity and diabetes are associated with the development of dementia in observational cohort studies^1^. If this association is causal, risk factors for vascular diseases may be responsible for around 40% of the cases of dementia worldwide^2^. However, the effect sizes seen in observational cohort studies are usually larger than those seen in randomized trials of vascular risk intervention to reduce cognitive decline or dementia^3^. This may be because cohort studies are limited by residual confounding, reverse causation, differential loss to follow-up, or selection biases^4,5^. More data from less biased designs are needed to triangulate the causal effects of vascular risk factors on the development of dementia.

Mendelian randomization (MR) uses genetic variants as proxies, or instrumental variables (IVs), to estimate a causal effect of an exposure on an outcome. MR is less susceptible to confounding and reverse causation than observational studies because genetic variants are assumed to be randomly assigned at meiosis^6^. As such, MR can be thought of as a “natural” randomized control trial. MR studies are subject to different biases from observational studies. Their validity rests on three key assumptions: (i) the genetic variant has a known association with the risk factor of interest; (ii) the genetic variant is not associated with a known confounder; and (iii) the genetic variant affects the outcome only through the risk factor of interest. As most genetic instrumental variables are only modestly associated with their exposures of interest, MR gives an unbiased but imprecise estimate^7^. Meta-analysis of MR studies could mitigate this imprecision. This approach has previously refined estimates of the effect obesity on vascular diseases^8^.

In this study, we meta-analysed MR studies of the association of modifiable vascular risk factors with dementia. Secondly, we estimated the heterogeneity between estimates from different MR studies for a given risk factor that used the same outcome cohort. Thirdly, we compared our meta-analysis estimates with estimates obtained from meta-analysis of observational studies.

## Methods

We used the Preferred Reporting Items for Systematic Reviews and Meta-Analyses (PRISMA) guideline to report this study^9^. A protocol has been developed and made available online (Supplementary Material 3)^10^. Minor amendments in study methodology have been made since its publication. Specifically, 1) the quality assessment questionnaire was shortened from 11 questions to 10 questions; 2) we applied a Bonferroni correction to account for the assessment of multiple risk factors; 3) MR meta-analysis results were compared with observational studies. No ethical approval was required.

### Search Strategy

We searched on OVID, Scopus and Web of Science, covering 13 databases: Medline, Embase, AMED, PsycINFO, BIOSIS Citation Index, Web of Science core collection, Current Contents Connect, Data Citation Index, Derwent Innovations Index, KCI-Korean Journal Database, Russian Science Citation Index, SciELO Citation Index, and Zoological Record. The search looked for a combination of dementia, and Mendelian randomization (Supplementary Material 1, Table 1). No risk factors were specified in the search query. We forward searched by screening for all referenced articles in the retrieved articles using Google Scholar. The final search was performed on 22^nd^ October 2021.

**Table 1.**
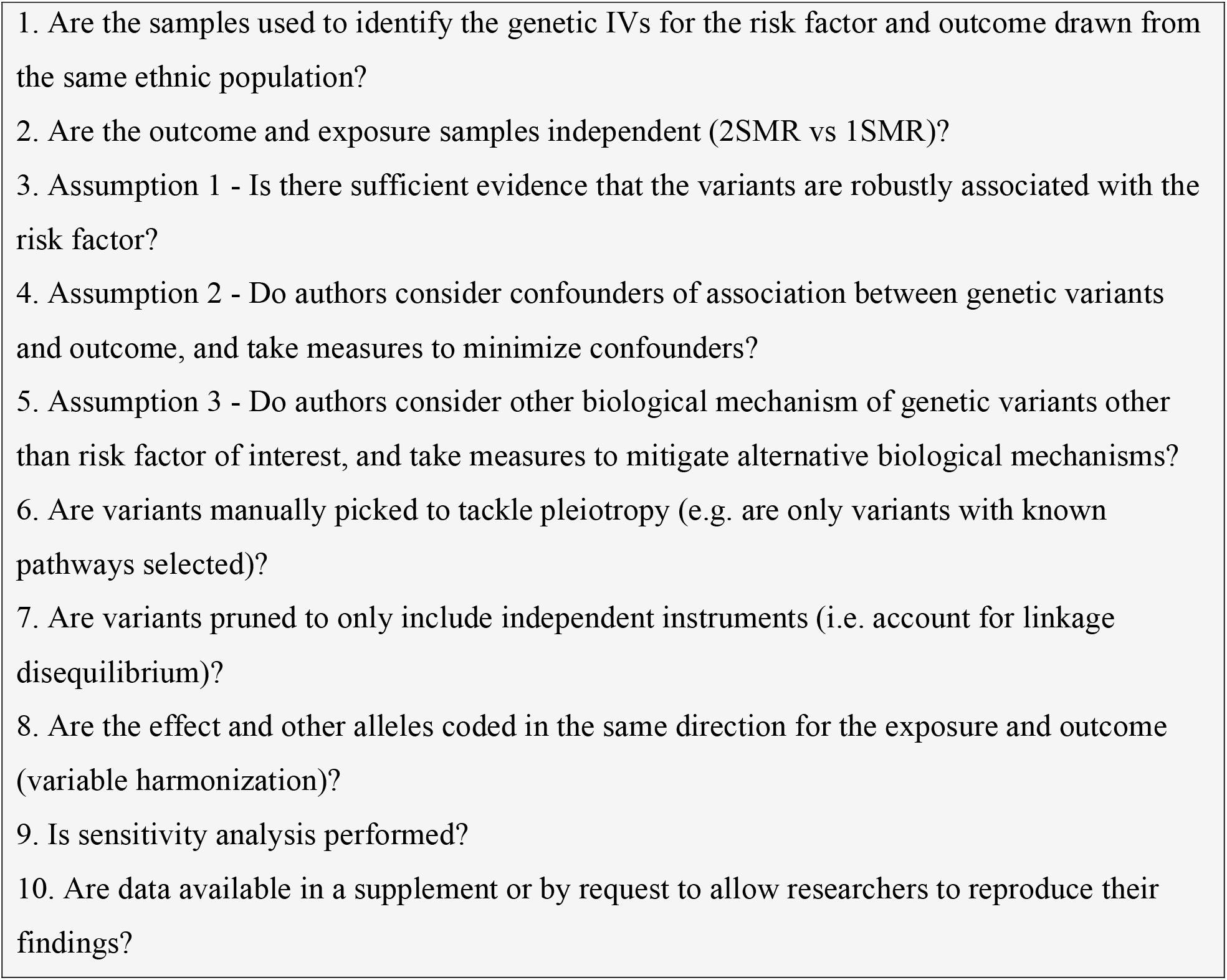
Quality assessment questionnaire. 2SMR = 2-sample Mendelian Randomization, has independent outcome and exposure samples. 1SMR = 1-sample Mendelian Randomization.

### Study selection

We included published or pre-print studies that used inverse-variance weighted (IVW) two-sample MR with a poly- or oligo-genetic instrument for type 2 diabetes mellitus, low density lipoprotein (LDL) cholesterol, high density lipoprotein (HDL) cholesterol, total cholesterol, triglycerides, systolic blood pressure, diastolic blood pressure, body mass index (BMI), or plasma glucose. Included studies reported a causal estimate value with an odds ratio (OR), hazard ratio (HR), risk ratio (RR) or β-coefficient by an absolute value of per unit increase, and associated 95% confidence interval (CI) or standard error. When interquartile range was reported, we estimated the standard deviation as interquartile range/1.35^11^.

We excluded studies that were duplicates, not written in English, or where no full text was available. We included only one estimate from each cohort per risk factor. Where more than one study had been carried out using a cohort, we included the highest quality study; where the quality of the studies was similar, we included the study with the most recent exposure GWAS. The outcomes were all cause dementia or late-onset Alzheimer’s disease (LOAD). Uncertainties were resolved by discussion with two other reviewers (WNW and RMW).

### Quality assessment

A quality assessment tool was developed by synthesizing three published guidelines for assessing MR studies^12–14^ (Table 1). Studies that did not satisfactorily address items 3-5, which describe the three core assumptions of MR, were excluded.

### Data extraction and meta-analysis

For each study, we extracted: GWAS source, ethnicity, number of single nucleotide polymorphisms (SNPs) used as instrumental variables for each risk factor, case/control sample size, effect estimates and units, and measures of variation were extracted for each study. When multiple analysis approaches were used to generate an effect estimate, the value generated using the most SNPs without compromising pleiotropy was used (linkage disequilibrium r^2^ < 0.2). Effect estimates that included IVs mapping to the *APOE* locus, which has a known association with dementia were excluded. For studies with missing data, the authors were emailed twice and the study was excluded if no reply was received. The full details are available in Supplementary Material 2, S1 & S2.

Effect estimates and measures of variation were standardized into common units for each risk factor. The effect estimates were pooled using a fixed effects model to generate an overall estimate for each risk factor of interest. Cochrane Q and I^2^ statistics were used to assess heterogeneity across studies. Analyses and plots were executed with the *Metafor* package (version 2.4-0) in R (version 4.0.3)^15^. A Bonferroni correction was applied to maintain a 5% family-wise error rate, yielding a significance threshold of 0.05 divided by n risk factors assessed (*p* = 0.05/9).

We performed sensitivity analysis by meta-analysing using alternative eligible studies, which were excluded in our primary meta-analysis due to outcome cohort overlap or the use of a superseded exposure GWAS. We substituted the studies with IGAP (2013) as the outcome cohort, with another study with the same outcome cohort with either the lowest or highest estimate in attempt to determine any significant change in overall effect estimate. Statistical significance was determined by applying a Bonferroni correction, as described above.

### Observational Study Comparison

#### Search Strategy

We searched on OVID, Scopus and Web of Science for a meta-analyses of cohort studies estimating the association between vascular risk factors with later dementia (Supplementary Material 1, Table 2).

**Table 2.** Quality assessment of studies included for meta-analysis. Yes = Reported, No = Not reported. Each question corresponds to the questions outlined in Table 1. *Ware *et al*. (2021) conducted a one sample MR.

|  | 16 | 17 | 18 | 19 | 20 | 21 | 22 | 23 | 24 | 25 | 26 | 27 |
| --- | --- | --- | --- | --- | --- | --- | --- | --- | --- | --- | --- | --- |
| **1. Are the samples used to identify the genetic IVs for the risk factor and outcome drawn from the same ethnic population?** | Y | Y | Y | Y | Y | Y | Y | Y | Y | Y | Y | Y |
| **2. Are the outcome and exposure samples independent (2SMR)?** | Y | Y | N* | Y | Y | Y | Y | Y | Y | Y | Y | Y |
| **3. Assumption 1 - Is there sufficient evidence that the variants are robustly associated with the risk factor?** | Y | Y | Y | Y | Y | Y | Y | Y | Y | Y | Y | Y |
| **4. Assumption 2 - Do authors consider confounders of association between genetic variants and outcome, and take measures to minimize confounders?** | Y | Y | Y | Y | Y | Y | Y | Y | Y | Y | Y | Y |
| **5. Assumption 3 - Do authors consider other biological mechanism of genetic variants other than risk factor of interest, and take measures to mitigate alternative biological mechanisms?** | Y | Y | Y | Y | Y | Y | Y | Y | Y | Y | Y | Y |
| **6. Are variants manually picked only with known mechanism?** | N | N | N | N | N | N | N | N | N | N | N | N |
| **7. Are variants pruned to only include independent instruments (i.e. account for linkage disequilibrium)?** | N | Y | N | Y | Y | N | Y | Y | Y | Y | Y | Y |
| **8. Are the effect and other alleles coded in the same direction for the exposure and outcome (variable harmonization)?** | Y | Y | N | N | N | N | N | N | Y | Y | N | N |
| **9. Is sensitivity analysis performed?** | Y | Y | Y | Y | Y | Y | Y | Y | Y | Y | Y | Y |
| **10. Are data available in a supplement or by request to allow researchers to reproduce their findings?** | Y | Y | Y | Y | Y | Y | Y | Y | Y | Y | Y | Y |

#### Study selection

We selected one representative meta-analysis of observational studies for each risk factor. We considered all articles in English which analysed cohorts, reported OR/HR/RR, and associated 95% confidence interval (CI) or standard error. If multiple meta-analyses were eligible for inclusion per given risk factor, we selected the study with the highest total number of participants.

## Results

### Literature Search

A search of three databases, OVID, Scopus and Web of Science, found 5211 unique articles. After applying inclusion and exclusion criteria to the abstracts, 29 articles were retained (Figure 1). Seventeen articles were not included for meta-analysis after full text review because either the results were not reported per unit increase in risk factor or there was an overlap of between outcome cohorts. Of those seventeen articles, ten studies were reserved for secondary outcome analyses.

**Figure 1.**
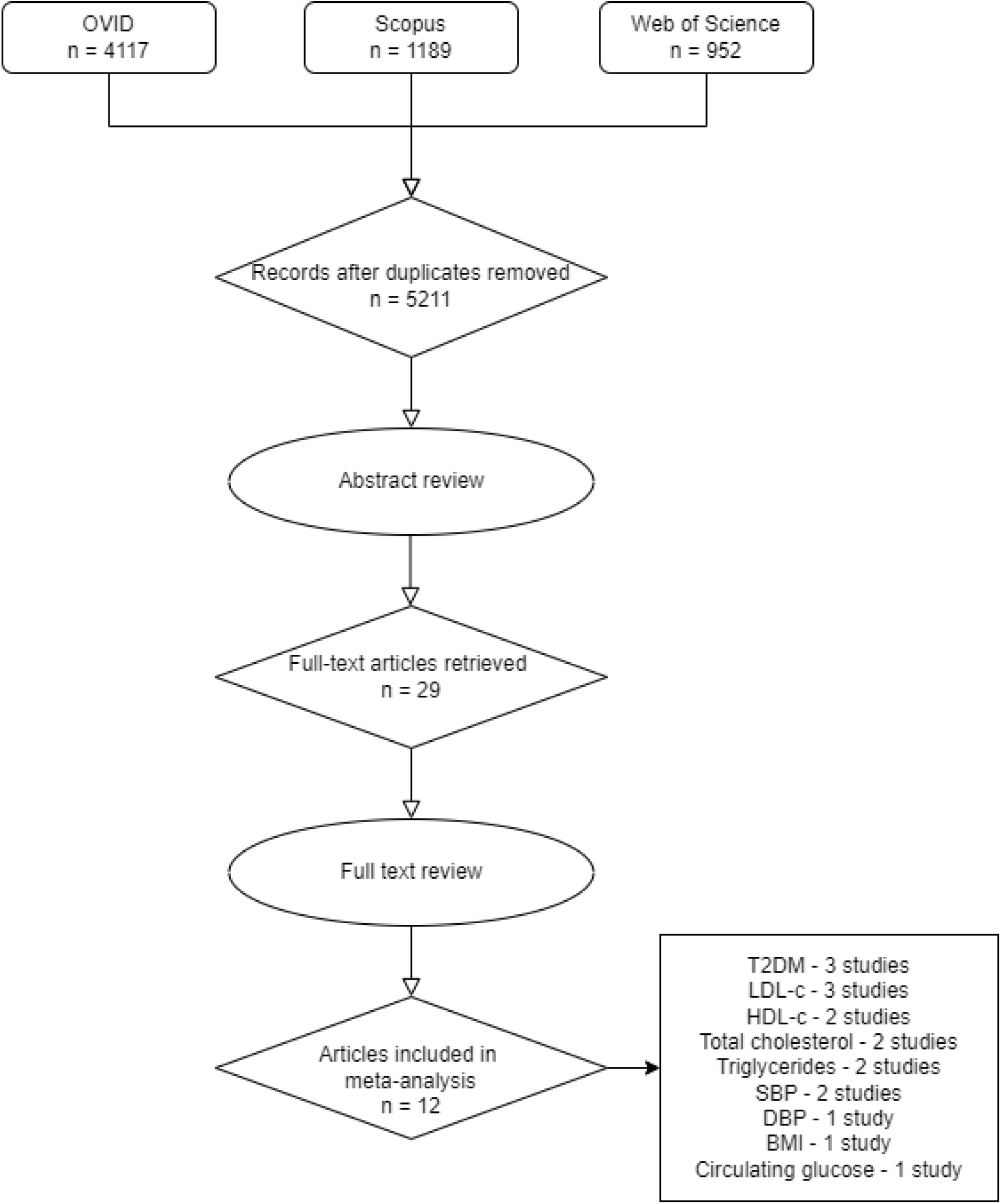
PRISMA flow diagram of the literature search results. Article relevance was assessed from title and abstract. Subsequently, full text was read for confirmation or further exclusion and additional studies were identified by forward searching from selected articles.

### Identification of primary MR studies

There were three primary MR studies each for type 2 diabetes^16–18^ and LDL cholesterol^19–21^; two studies each for HDL cholesterol^19,22^, total cholesterol^19,23^, triglycerides^19,21^, and systolic blood pressure^20,24^; and one study each for diastolic blood pressure^25^, BMI^26^ and circulating glucose^27^ (Figure 1). The MR studies for diastolic blood pressure, BMI, and circulating glucose reported an overall estimate produced through the meta-analysis of two or more outcome cohorts. As such, the estimates from these studies were included in the present study.

### Mendelian Randomization Meta-Analysis

Quality assessment was performed on the 12 selected studies (Table 2). All studies addressed the three assumptions of MR. The MR results for each of the six vascular risk factors with more than one MR study were meta-analysed (Figure 2). BMI was significantly associated with dementia as stated in the original study by Li *et al*. (2021), with higher BMI increasing the odds of developing dementia (1.03 [1.01, 1.05] per 5 kg/m^2^ increase, p = 0.00285), and met the corrected significance threshold.

**Figure 2.**
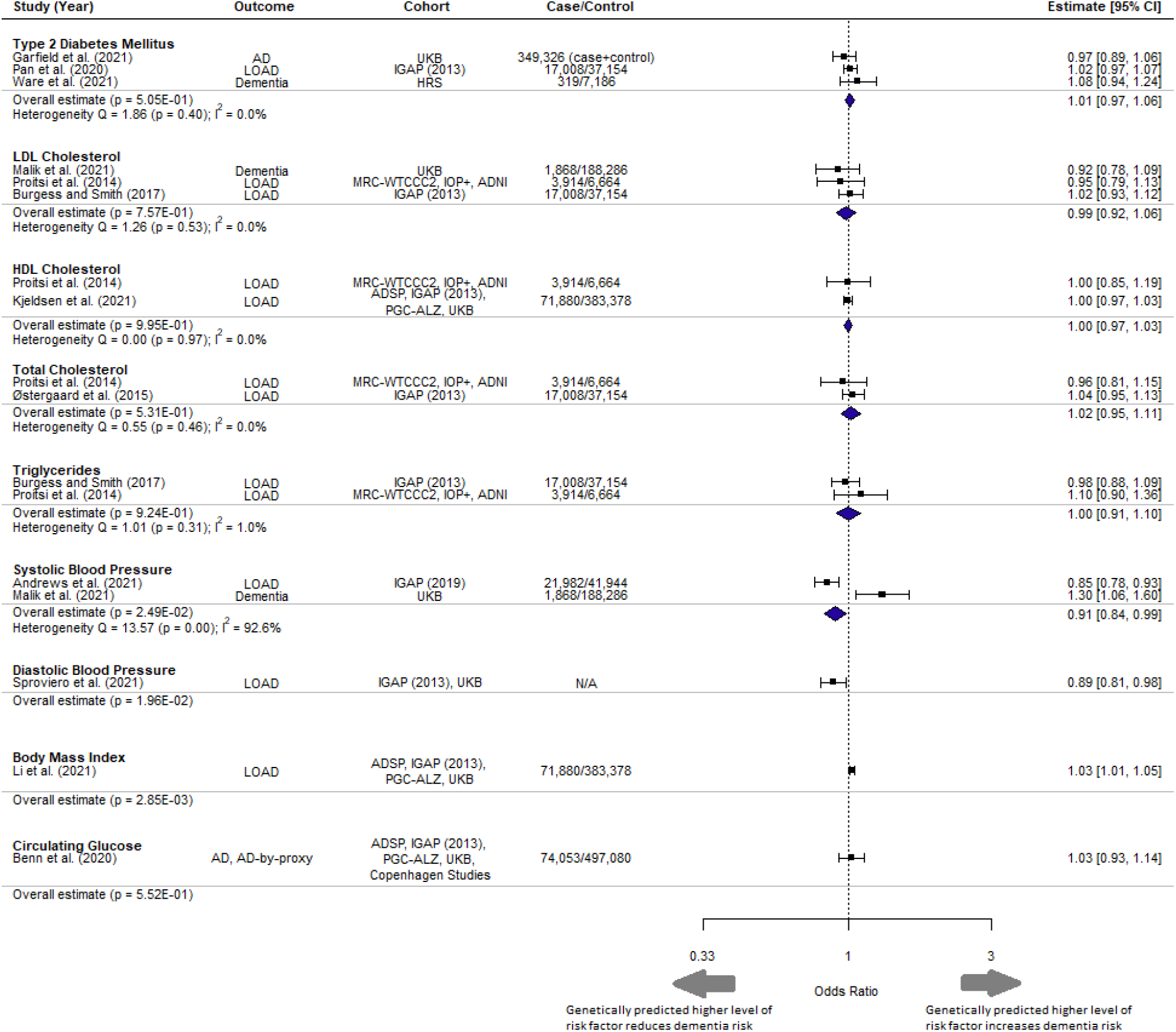
Forest plot of meta-analysis results with overall estimate in OR, 95% CI and p-value for each exposure. UKB = UK Biobank, IGAP = International Genomics of Alzheimer’s Project, HRS = Health and Retirement Study, MRC-WTCCC2 = Medical Research Council (MRC)-Wellcome Trust Case Control Consortium, IOP+ = Institute of Psychiatry Plus, ADNI = Alzheimer’s Disease Neuroimaging Initiative. Copenhagen Studies = Copenhagen General Population Study and the Copenhagen City Heart Study.

Comparison of different MR studies using the same cohort (IGAP) had similar estimates (I^2^ = 0%) for all risk factors, apart from for LDL-c (I^2^ = 65.2%) and systolic blood pressure (I^2^ = 25.2%) (Figure 3). The MR studies included in this analysis all fulfilled the three core assumptions of MR. Sensitivity analysis to replace MR studies using the IGAP (2013) outcome GWAS with another MR study with the most extreme values rendered the meta-analysed effect estimate for all risk factors to remain non-significant (Supplementary Material 1, Figure 1). Diastolic blood pressure, BMI and circulating glucose were excluded from sensitivity analysis because only a single study was reported for each risk factor in this study.

**Figure 3.**
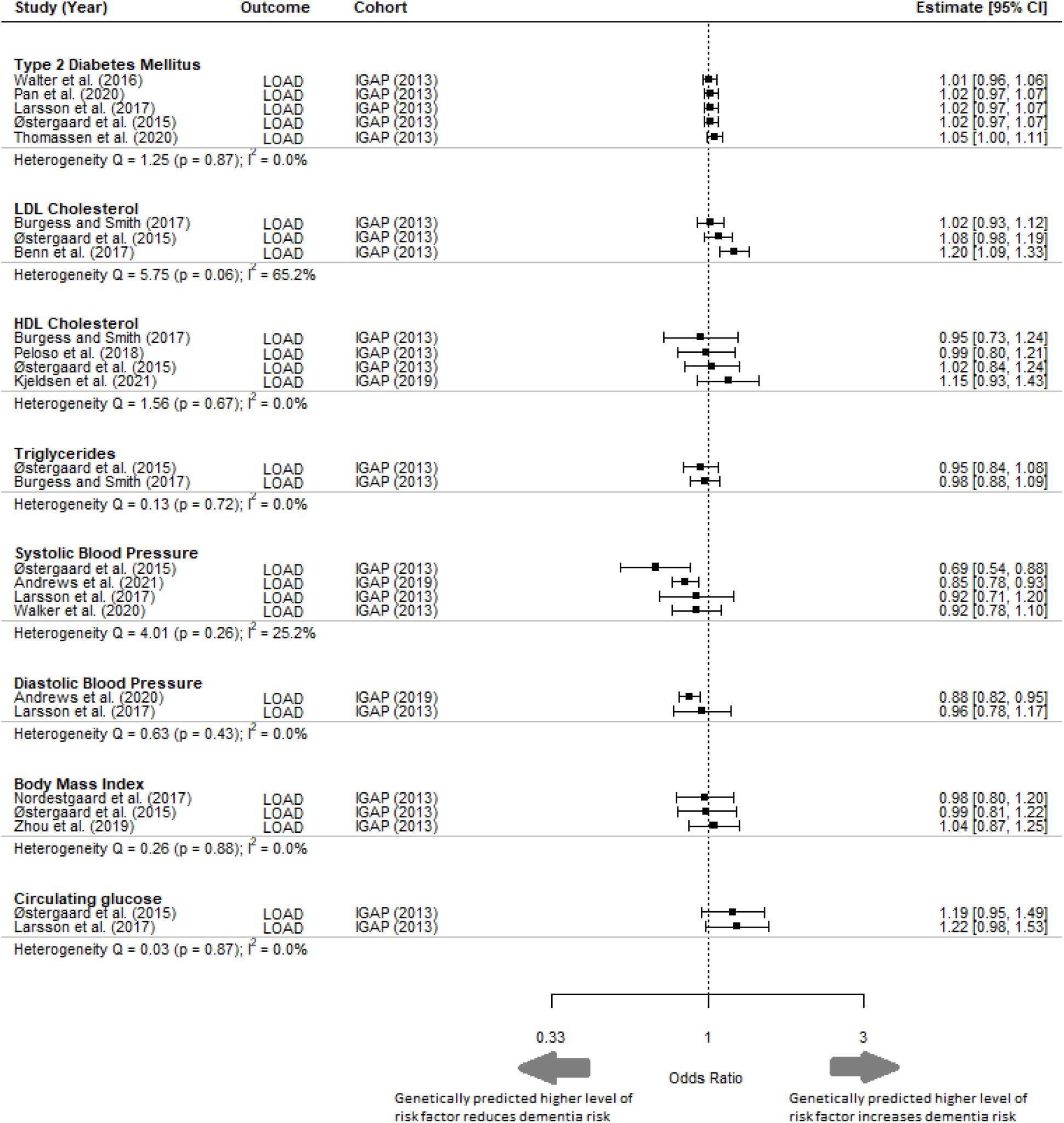
Forest plot of primary MR studies with same outcome cohort IGAP (2013). This figure includes studies that have not been selected in the meta-analysis and is shown to highlight the heterogeneity of results despite using the same outcome cohort. LOAD = Late onset Alzheimer’s disease; IGAP = International Genomics of Alzheimer’s Project; CI = Confidence Interval.

### Observational Study Comparison

For eight of the risk factors, we compared the effect estimate from MR studies with the effect estimate from the largest available meta-analysis of observational studies (Figure 4) ^28–33^. One risk factor, circulating glucose, did not have an eligible meta-analysis; therefore, the primary study with largest study cohort was included as the comparator^34^. The units of estimates from most meta-analyses of observational studies were not explicitly stated, so it was not possible to compare the magnitude of effect with that obtained from the meta-analysis of MR studies. There were significant associations between LDL cholesterol, diastolic blood pressure diabetes and circulating glucose with a higher risk of later dementia in observational studies, but neutral associations from studies using MR.

**Figure 4.**
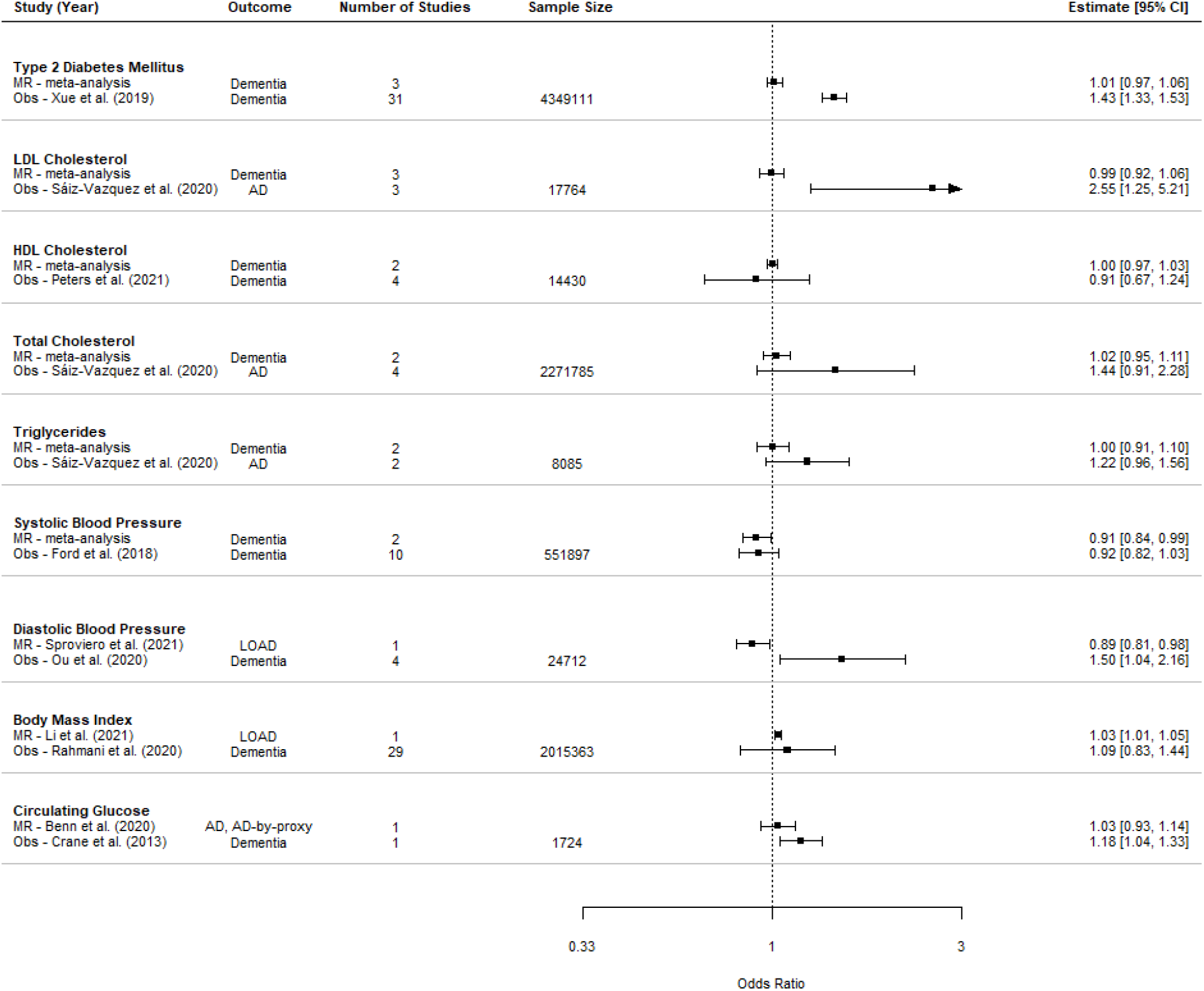
Comparison of estimates from meta-analysis of MR and observational studies (Obs). Sample size is the sum of control and case numbers.

## Discussion

We performed a meta-analysis of MR studies assessing the effects of nine modifiable risk factors for vascular diseases on the odds of dementia. With the exception of for BMI, we did not obtain evidence for an association between genetically predicted levels of any vascular risk factor and the odds of developing dementia.

Fewer vascular risk factors were associated with dementia based on meta-analysed estimates from MR studies than from meta-analyses of observational studies. This may be because the estimates from cohort studies were limited by residual confounding, or by a selection bias that identified populations with particularly high risk of dementia due to vascular risk factors, or by publication bias towards reporting positive results. The MR studies may have been limited by weak genetic instruments that explained only a small percentage of the variation in the risk factors of interest. For example, in Andrews et al. (2021) and Malik et al. (2021) less than 6% of the variation in blood pressure was explained by all the SNPs identified through GWASs. Moreover, these MR studies are unable to distinguish age-dependent mechanisms. For instance, high systolic blood pressure has been linked with harmful effects during mid-life but protective effects during late-life (>75)^35,36^. A similar case has been made for cholesterol levels^37^. There is additional uncertainty regarding the reliability of GWASs themselves in which genetic instruments were derived from. GWASs may have been underpowered, resulting in unreliable identification of IV SNPs^38^. The populations included in the GWASs for exposure and outcome may differ, limiting their comparability. An attempt to mitigate this issue was, however, made by using GWAS studies that focused on populations of European descent. A further consideration is that differences in model formulation between the GWASs from which IVs are identified and observational studies has the potential to limit the comparability of the phenotype under consideration.

An exploration of the consistency between the findings of MR studies that used different exposure GWASs but the same outcome GWAS (citation) found consistency between the MR estimates for all risk factors, except for LDL cholesterol and systolic blood pressure. This may be because of differences in the SNPs used as IVs for these risk factors. Sometimes differences in the SNPs used as IVs are attributable to a decision to focus on specific pathways: for example, Benn *et al*. (2017) focused on *PCSK9* and *HMGCR* variants for LDL cholesterol to tackle pathways that are therapeutically relevant based on PCSK9 inhibitors and statins^39^. They reported a statistically significant causative effect of LDL-c, unlike other studies that included SNPs from other genomic regions (Figure 3). However, the choice of SNPs may also be simply due to differences in available data. For instance, Østergaard et al. (2015) obtained 24 SNPs to act as IVs for systolic blood pressure from an up-to-date GWAS at that time, while Larsson et al. (2021) obtained 93 IV SNPs from a study published in 2017. Østergaard et al. (2015) concluded that higher systolic blood pressure lowered the odds of dementia, while Larsson et al. (2021) found no significant association (Figure 3). Therefore, recognizing these differences is important when interpreting the estimates from MR studies.

### Limitations

We identified relatively few eligible studies. Although many potentially relevant primary MR studies were identified, many studies used data from the same source; for example, many studies of Alzheimer’s disease use IGAP (2013)^40^. Therefore, many otherwise eligible studies had to be excluded to ensure independence of estimates in our meta-analysis (Supplementary Material 2, S2). The limited number of eligible studies led us to pool our two outcomes of interest (Alzheimer’s disease and dementia). Although Alzheimer’s disease constitutes 60-80% of dementia cases, a significant proportion of dementia cases are associated with different diseases with distinct pathophysiology such as vascular dementia^41^.

Discrepancies in pathophysiology may potentially be reflected in the heterogeneity of estimates as seen in systolic blood pressure where outcomes Alzheimer’s disease and dementia were both present (I^2^ = 92.6%) (Figure 2).

Secondly, although we meta-analysed studies with unique outcome cohorts, we did not have access to individual participant data. We were therefore unable to control any potential overlap between the cohorts.

Thirdly, we found heterogeneity between the MR estimates obtained using the same outcome cohort for two risk factors, systolic blood pressure and LDL cholesterol. This was in spite of key similarities in the study, such as only including population of European ancestry and using the same GWAS to derive the SNPs. The heterogeneity observed is likely to stem from differences in methods, such as discrepancy in rationale for selection of SNPs, p-value cutoff for the use of SNPs as IVs, and discrepancy in covariates when analysing exposure-outcome relationship. The exact effect these differences have on the estimate and its clinical implications remains yet to be characterized. There is a need to assess the effect of such discrepancies and the robustness of MR estimates through sensitivity analyses.

## Conclusion

Out of the nine vascular risk factors assessed in this study, only genetically predicted BMI showed evidence of being causally associated with dementia. Estimates from observational studies for many risk factors were significantly associated with dementia.

## Data Availability

All data produced in the present study are available upon reasonable request to the authors.

## References

1. Baumgart M, Snyder HM, Carrillo MC, Fazio S, Kim H, Johns H. Summary of the evidence on modifiable risk factors for cognitive decline and dementia: A population-based perspective. Alzheimer’s & Dementia. 2015;11(6):718–726. doi:10.1016/J.JALZ.2015.05.016

2. Livingston G, Huntley J, Sommerlad A, et al. Dementia prevention, intervention, and care: 2020 report of the Lancet Commission. The Lancet. 2020;396(10248):413–446. doi:10.1016/S0140-6736(20)30367-6

3. Hughes D, Judge C, Murphy R, et al. Association of Blood Pressure Lowering With Incident Dementia or Cognitive Impairment: A Systematic Review and Meta-analysis. JAMA. 2020;323(19):1. doi:10.1001/JAMA.2020.4249

4. Patel CJ, Burford B, Ioannidis JPA. Assessment of vibration of effects due to model specification can demonstrate the instability of observational associations. Journal of clinical epidemiology. 2015;68(9):1046. doi:10.1016/J.JCLINEPI.2015.05.029

5. Carlson MDA, Morrison RS. Study design, precision, and validity in observational studies. Journal of Palliative Medicine. 2009;12(1):77–82. doi:10.1089/jpm.2008.9690

6. Smith GD, Hemani G. Mendelian randomization: Genetic anchors for causal inference in epidemiological studies. Human Molecular Genetics. 2014;23(R1). doi:10.1093/hmg/ddu328

7. Burgess S, Thompson SG, Collaboration CCG. Avoiding bias from weak instruments in Mendelian randomization studies. International Journal of Epidemiology. 2011;40(3):755–764. doi:10.1093/IJE/DYR036

8. Riaz H, Khan MS, Siddiqi TJ, et al. Association Between Obesity and Cardiovascular Outcomes: A Systematic Review and Meta-analysis of Mendelian Randomization Studies. JAMA Network Open. 2018;1(7):e183788. doi:10.1001/JAMANETWORKOPEN.2018.3788

9. Moher D, Liberati A, Tetzlaff J, et al. Preferred reporting items for systematic reviews and meta-analyses: The PRISMA statement. PLoS Medicine. 2009;6(7). doi:10.1371/journal.pmed.1000097

10. Systematic review and meta-analysis of Mendelian randomisation studies on modifiable risk factors for dementia. Accessed February 16, 2022. https://www.protocols.io/view/systematic-review-and-meta-analysis-of-mendelian-r-bpeemjbe

11. Wan X, Wang W, Liu J, Tong T. Estimating the sample mean and standard deviation from the sample size, median, range and/or interquartile range. BMC medical research methodology. 2014;14(1). doi:10.1186/1471-2288-14-135

12. Davies NM, Holmes M V., Davey Smith G. Reading Mendelian randomisation studies: A guide, glossary, and checklist for clinicians. BMJ (Online). 2018;362. doi:10.1136/bmj.k601

13. Grover S, Del Greco F, König IR. Evaluating the current state of Mendelian randomization studies: A protocol for a systematic review on methodological and clinical aspects using neurodegenerative disorders as outcome. Systematic Reviews. 2018;7(1):145. doi:10.1186/s13643-018-0809-3

14. Burgess S, Davey Smith G, Davies NM, et al. Guidelines for performing Mendelian randomization investigations. Wellcome Open Research. 2020;4. doi:10.12688/wellcomeopenres.15555.2

15. Viechtbauer W. Conducting meta-analyses in R with the metafor. Journal of Statistical Software. 2010;36(3). doi:10.18637/jss.v036.i03

16. Garfield V, Farmaki AE, Fatemifar G, et al. The Relationship Between Glycaemia, Cognitive Function, Structural Brain Outcomes and Dementia: A Mendelian Randomisation Study in the UK Biobank. Diabetes. Published online June 15, 2021:db200895. doi:10.2337/DB20-0895

17. Pan Y, Chen W, Yan H, Wang M, Xiang X. Glycemic traits and Alzheimer’s disease: a Mendelian randomization study. Aging (Albany NY). 2020;12(22):22688. doi:10.18632/AGING.103887

18. Ware EB, Morataya C, Fu M, Bakulski KM. Type 2 Diabetes and Cognitive Status in the Health and Retirement Study: A Mendelian Randomization Approach. Frontiers in Genetics. 2021;12:634767. doi:10.3389/FGENE.2021.634767

19. Proitsi P, Lupton MK, Velayudhan L, et al. Genetic Predisposition to Increased Blood Cholesterol and Triglyceride Lipid Levels and Risk of Alzheimer Disease: A Mendelian Randomization Analysis. PLoS Medicine. 2014;11(9). doi:10.1371/JOURNAL.PMED.1001713

20. Malik R, Georgakis MK, Neitzel J, et al. Midlife vascular risk factors and risk of incident dementia: Longitudinal cohort and Mendelian randomization analyses in the UK Biobank. Alzheimer’s & Dementia. Published online 2021. doi:10.1002/ALZ.12320

21. Burgess S, Smith GD. Mendelian Randomization Implicates High-Density Lipoprotein Cholesterol– Associated Mechanisms in Etiology of Age-Related Macular Degeneration. Ophthalmology. 2017;124(8):1165. doi:10.1016/J.OPHTHA.2017.03.042

22. Kjeldsen EW, Thomassen JQ, Juul Rasmussen I, Nordestgaard BG, Tybjærg-Hansen A, Frikke-Schmidt R. Plasma HDL cholesterol and risk of dementia - observational and genetic studies. Cardiovascular research. Published online May 8, 2021. doi:10.1093/CVR/CVAB164

23. Østergaard SD, Mukherjee S, Sharp SJ, et al. Associations between Potentially Modifiable Risk Factors and Alzheimer Disease: A Mendelian Randomization Study. PLOS Medicine. 2015;12(6):e1001841. doi:10.1371/JOURNAL.PMED.1001841

24. Andrews SJ, Fulton-Howard B, O’Reilly P, Marcora E, Goate AM, Consortium collaborators of the ADG. Causal Associations Between Modifiable Risk Factors and the Alzheimer’s Phenome. Annals of neurology. 2021;89(1):54. doi:10.1002/ANA.25918

25. Sproviero W, Winchester L, Newby D, et al. High Blood Pressure and Risk of Dementia: A Two-Sample Mendelian Randomization Study in the UK Biobank. Biological psychiatry. 2021;89(8):817–824. doi:10.1016/J.BIOPSYCH.2020.12.015

26. Li X, Tian Y, Yang YX, et al. Life Course Adiposity and Alzheimer’s Disease: A Mendelian Randomization Study. Journal of Alzheimer’s disease : JAD. 2021;82(2):503–512. doi:10.3233/JAD-210345

27. Benn M, Nordestgaard BG, Tybjærg-Hansen A, Frikke-Schmidt R. Impact of glucose on risk of dementia: Mendelian randomisation studies in 115,875 individuals. Diabetologia 2020 63:6. 2020;63(6):1151–1161. doi:10.1007/S00125-020-05124-5

28. Rahmani J, Roudsari AH, Bawadi H, et al. Body mass index and risk of Parkinson, Alzheimer, Dementia, and Dementia mortality: a systematic review and dose-response meta-analysis of cohort studies among 5 million participants. Nutritional neuroscience. Published online 2020. doi:10.1080/1028415X.2020.1758888

29. Xue M, Xu W, Ou YN, et al. Diabetes mellitus and risks of cognitive impairment and dementia: A systematic review and meta-analysis of 144 prospective studies. Ageing research reviews. 2019;55. doi:10.1016/J.ARR.2019.100944

30. Sáiz-Vazquez O, Puente-Martínez A, Ubillos-Landa S, Pacheco-Bonrostro J, Santabárbara J. Cholesterol and Alzheimer’s disease risk: A meta-meta-analysis. Brain Sciences. 2020;10(6):1–13. doi:10.3390/brainsci10060386

31. Peters R, Xu Y, Antikainen R, et al. Evaluation of High Cholesterol and Risk of Dementia and Cognitive Decline in Older Adults Using Individual Patient Meta-Analysis. Dementia and geriatric cognitive disorders. 2021;50(4). doi:10.1159/000519452

32. Ford E, Greenslade N, Paudyal P, et al. Predicting dementia from primary care records: A systematic review and meta-analysis. PLoS ONE. 2018;13(3). doi:10.1371/JOURNAL.PONE.0194735

33. Ou YN, Tan CC, Shen XN, et al. Blood Pressure and Risks of Cognitive Impairment and Dementia: A Systematic Review and Meta-Analysis of 209 Prospective Studies. Hypertension. 2020;76(1):217–225. doi:10.1161/HYPERTENSIONAHA.120.14993

34. Crane PK, Walker R, Hubbard RA, et al. Glucose levels and risk of dementia. Forschende Komplementarmedizin. 2013;20(5):386–387. doi:10.1056/NEJMOA1215740/SUPPL_FILE/NEJMOA1215740_DISCLOSURES.PDF

35. M K, EL H, MP L, et al. Midlife vascular risk factors and Alzheimer’s disease in later life: longitudinal, population based study. BMJ (Clinical research ed). 2001;322(7300):1447–1451. doi:10.1136/BMJ.322.7300.1447

36. Qiu C, Strauss E von, Winblad B, Fratiglioni L. Decline in Blood Pressure Over Time and Risk of Dementia. Stroke. 2004;35(8):1810–1815. doi:10.1161/01.STR.0000133128.42462.EF

37. van Vliet P. Cholesterol and late-life cognitive decline. Journal of Alzheimer’s disease : JAD. 2012;30 Suppl 2(SUPPL.2). doi:10.3233/JAD-2011-111028

38. Tam V, Patel N, Turcotte M, Bossé Y, Paré G, Meyre D. Benefits and limitations of genome-wide association studies. Nature Reviews Genetics 2019 20:8. 2019;20(8):467–484. doi:10.1038/s41576-019-0127-1

39. M. Benn, B. G. Nordestgaard, R. Frikke-Schmidt ATH. Low LDL cholesterol, PCSK9 and HMGCR genetic variation, and risk of Alzheimer’s disease and Parkinson’s disease: Mendelian randomisation study. BMJ. 2017;357. doi:10.1136/BMJ.J3170

40. Lambert JC, Ibrahim-Verbaas CA, Harold D, et al. Meta-analysis of 74,046 individuals identifies 11 new susceptibility loci for Alzheimer’s disease. Nature Genetics. 2013;45(12):1452–1458. doi:10.1038/ng.2802

41. 2021 Alzheimer’s disease facts and figures. Alzheimer’s and Dementia. 2021;17(3):327–406. doi:10.1002/alz.12328

