## Supplementary material 1 for "Assessing the role of vascular risk factors in dementia: Mendelian randomization meta-analysis and comparison with observational estimates"

**Supplementary Table 1**

| Database | Search terms | Articles retrieved |
|---|---|---|
| **Ovid** | (exp Dementia/ or exp Cognition Disorders/ or dement* or alzheimer* or cognit* or neurocognit* or ((cognit* or memory or cerebr* or mental*) adj3 (declin* or impair* or los* or deteriorat* or degenerat* or deficit* or complain* or disturb* or disorder* or dysfunction* or insufficien* or fail*)))<br>AND<br>(exp Mendelian Randomization Analysis/ or (Mendelian adj1 random*) or (MR not (magnetic or resonance or imag* or receptor)) or (instrumental adj1 variable*) or (genetic adj1 instrument)) | 4117<br>[final search conducted on 22nd October] |
| **Scopus** | TITLE-ABS-KEY (("dement*" OR "Alzheimer*" OR "cognit*" OR "neurocognit*" OR "memory" OR "cerebr*" OR "mental*")<br>AND<br>("Mendelian random*" OR "instrumental variable*" OR "genetic instrument*")) | 1189<br>[final search conducted on 22nd October] |
| **Web of Science** | TS=(dement* OR Alzheimer* OR cognit* OR neurocognit* OR memory OR cerebr* OR mental*)<br>AND<br>TS=(“Mendelian random*” OR “instrumental variable*” OR “genetic instrument*") | 952<br>[final search conducted on 22nd October] |

Supplementary Table 1. Search strategy terms. Search terms were different for each database in order to enter an acceptable syntax. Ovid and Web of Science collectively covered 14 databases: Medline, Embase, AMED, PsycINFO, Scopus database, BIOSIS Citation Index, Web of Science core collection, Current Contents Connect, Data Citation Index, Derwent Innovations Index, KCI-Korean Journal Database, Russian Science Citation Index, SciELO Citation Index, Zoological Record.

**Supplementary Table 2**

| Database | Search terms | Articles retrieved |
|---|---|---|
| **Ovid** | (exp Dementia/ or exp Cognition Disorders/ or dement* or alzheimer* or cognit* or neurocognit* or ((cognit* or memory or cerebr* or mental*) adj3 (declin* or impair* or los* or deteriorat* or degenerat* or deficit* or complain* or disturb* or disorder* or dysfunction* or insufficien* or fail*)))<br>AND<br>(exp Body Mass Index/ or exp Glucose/ or exp Diabetes Mellitus, Type 2/ or exp Cholesterol/ or exp Triglycerides/ or exp Blood Pressure/)<br>AND (exp Meta-analysis/ or meta) | 1937<br>[final search conducted on 27th November] |
| **Scopus** | TITLE-ABS-KEY (("dement*" OR "Alzheimer*")<br>AND<br>(“body mass index” OR “BMI” OR “diabetes” OR “glucose” OR “lipid*” OR “cholesterol” OR “triglycerides” OR “blood pressure”)<br>AND (“meta-analysis” OR “meta”)) | 1053<br>[final search conducted on 27th November] |
| **Web of Science** | TS=(dement* OR Alzheimer*)<br>AND<br>TS=(“body mass index” OR “BMI” OR “diabetes” OR “glucose” OR lipid* OR “cholesterol” OR “triglycerides” OR “blood pressure”)<br>AND TS=(meta-analys*) | 573<br>[final search conducted on 27th November] |

Supplementary Table 1. Search strategy terms. Search terms were different for each database in order to enter an acceptable syntax. Ovid and Web of Science collectively covered 5 databases: Medline, Embase, AMED, PsycINFO, Scopus database, BIOSIS Citation Index, Web of Science core collection, Current Contents Connect, Data Citation Index, Derwent Innovations Index, KCI-Korean Journal Database, Russian Science Citation Index, SciELO Citation Index, Zoological Record.

### Supplementary Figure

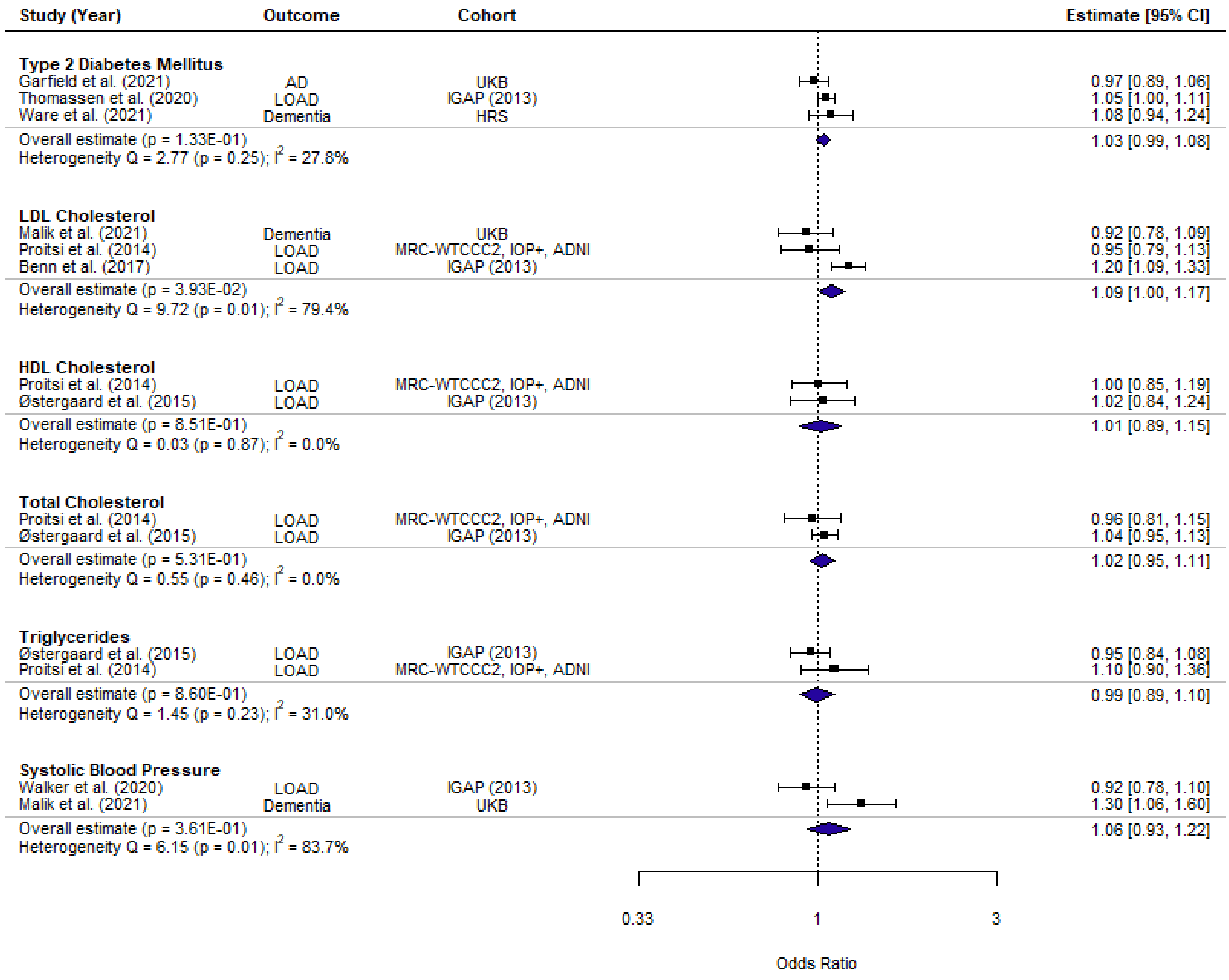


Supplementary Figure 2. Forest plot of primary MR studies for sensitivity analysis. Studies were artificially picked to maximize skewing in either direction. One study per unique outcome cohort was maintained. Diastolic blood pressure, BMI and circulating glucose were omitted due to no meta-analysis being performed as part of our main analysis. Bonferroni corrected p-value threshold = 0.008 (0.05/6).
